# Shared brain and genetic architectures between mental health and physical activity

**DOI:** 10.1101/2021.12.28.21268486

**Authors:** Wei Zhang, Sarah E. Paul, Anderson Winkler, Ryan Bogdan, Janine D. Bijsterbosch

## Abstract

Physical activity is correlated with, and effectively treats various forms of psychopathology. However, whether biological correlates of physical activity and psychopathology are shared remains unclear. Here, we examined the extent to which the neural and genetic architecture of physical activity and mental health are shared. Using data from the UK Biobank (N=6,389), canonical correlation analysis was applied to estimate associations between the amplitude and connectivity strength of sub-networks of three major neurocognitive networks (default mode, DMN; salience, SN; central executive networks, CEN) with accelerometer-derived measures of physical activity and self-reported mental health measures (primarily of depression, anxiety disorders, neuroticism, subjective well-being, and risk-taking behaviors). We estimated the genetic correlation between mental health and physical activity measures, as well as putative causal relationships by applying linkage disequilibrium score regression, genomic structural equational modeling, and latent causal variable analysis to genome-wide association summary statistics (GWAS N=91,105-500,199). Physical activity and mental health were associated with connectivity strength and amplitude of the DMN, SN, and CEN (r’s≥0.12, p’s<0.048). These neural correlates exhibited highly similar loading patterns across mental health and physical activity models even when accounting for their shared variance. This suggests a largely shared brain network architecture between mental health and physical activity. Mental health and physical activity (including sleep) were also genetically correlated (|rg|=0.085-0.121), but we found no evidence for causal relationships between them. Collectively, our findings provide empirical evidence that mental health and physical activity have shared brain and genetic architectures and suggest potential candidate sub-networks for future studies on brain mechanisms underlying beneficial effects of physical activity on mental health.

## 1. Introduction

Mental health and physical activity have both been linked to emotion, cognition, and brain correlates^1–4^. Furthermore, physical activity is known to improve various psychiatric conditions^5– 7^. Yet, it remains elusive whether mental health and physical activity overlap in neural and genetic architectures. Here, in a large population sample (n=6,389 from the UK Biobank ^8^), we estimate the extent to which physical activity and general mental health share patterns of resting-state functional MRI network measures and genetic architectures.

### 1.1 Potential overlap between mental health and physical activity in neural networks

Functional alterations in large-scale brain networks have been consistently implicated in a wide range of psychiatric disorders^9^. Dysfunctional configuration of neurocognitive networks such as the default mode, salience, and central executive networks has been hypothesized to characterize major psychiatric disorders including depression and anxiety (DMN, SN, CEN)^10^. In line with this hypothesis, findings from meta-analyses have shown that core cognitive and affective abnormalities in major depression can be accounted for by hypo-connectivity within the CEN and hyper-connectivity within the DMN^11^, together with hypo-connectivity between the control systems (i.e., CEN) and salience, emotion processing systems (i.e., SN)^12^. Similarly, a recent meta-analysis suggests that anxiety disorders are characterized by hypo-connectivity between subcortical limbic circuits that partially overlap with the SN, CEN, and DMN, as well as decoupling between the CEN and DMN^13^. Furthermore, the personality trait neuroticism, which is considered a risk marker for psychopathology, has also been linked to alterations in functional brain networks^14^. These same networks have also been linked to physical activity. Evidence from fMRI studies on physical activity demonstrated the changes in activity of and functional connectivity between these network hub regions including the hippocampus, parahippocampus, dorsal anterior cingulate cortex, and ventromedial prefrontal cortex that primarily subserve executive functions such as working memory, attention, and inhibition^15–19^. At the more system level, a 12-month aerobic walking intervention was found to increase resting-state functional connectivity between subnetworks of DMN, and between subnetworks of SN^20^, whereas connectivity in the CEN was found to increase after multiple sessions of high intensity interval training^21^. Interestingly, the intensity of physical exercise appeared to modulate functional connectivity changes in the hub regions of the CEN^22^, as well as in the DMN subsystems acutely and after 3 months of training^23^. Additionally, although sleep might not be considered a type of physical activity for its inactive nature, it is closely connected to mental health such that sleep problems have been considered as a risk factor for subsequent development of depressive symptoms^24,25^, and disrupted sleep is often seen in patients with various types of mood and anxiety disorders^26–28^.

Additionally, sleep disturbance can reduce physical activity levels and increase the risk of exercise-related injuries^29^. Although sleep has been largely overlooked in the literature for physical activity in relation to mental health, we included it in the current study for its close relationships with both mental health and physical activity. Sleep duration can be accurately measured with ecological sampling approaches such as wrist-worn accelerometers, which have been adopted in medical and mental health research^30,31^. At the neural network level, sleep deprivation has been linked to reduction in functional connectivity within the DMN ^32–34^, whereas increased sleep duration has been linked to strengthened connectivity within the DMN but reduced connectivity between DMN and SN^35^. Fluctuations in arousal (i.e., indication of drowsiness) during resting-state scan acquisitions have also been linked to the fMRI signal amplitude in sensorimotor networks^36^. These studies together highlight the engagement of the DMN, SN and CEN in both mental health and physical activity and demonstrate that the interplays between these large-scale intrinsic networks and subnetworks are coupled with changes in mental health symptoms and physical exercises. Yet, it remains unclear whether similar connectivity patterns or signal changes of these networks are shared by mental health and physical activity.

### 1.2 Potential overlap between mental health and physical activity in genetic variance

Physical activity is known to promote resilience to various psychiatric conditions, alleviating symptoms of depression, anxiety, and negative mood^37,38^. This buffering effect may be rooted in the shared genetic variance between mental health and physical activity. For instance, recent research employing summary statistics from independent genome-wide associate studies (GWAS) showed that higher polygenic risk scores for depression are associated with increased odds of incident depression, whereas self-reported physical activities such as walking, jogging, running, dancing and yoga appeared to reduce the odds with similar magnitude ^39^. This effect has also been observed using more objective measurement of physical activity such that reduced activity levels measured by accelerometer were found to associate with diagnoses of schizophrenia, bipolar disorder, depression, or autism spectrum disorders (ASD) and that healthy participants without disorder diagnoses were observed to perform less physical activity if they had a higher polygenic risk score f or schizophrenia, depression, and ASD ^40^.

Additionally, overlapping genetic architectures may exist across various psychiatric disorders including anxiety and depression^41–43^ as psychiatric phenotypes are highly polygenic ^44^. Thus, latent genetic factors capturing shared variance across clusters of psychiatric symptoms may improve identification of associations between mental health and physical activity. For instance, genomic contributions to disorders such as depression and anxiety are captured by a genetic factor for internalizing disorders that are primarily characterized by these two disorders as indicated by a confirmatory factor analysis, and this genetic factor is positively genetically associated with various adverse health outcomes and negatively genetically associated with physical movement patterns^45^. These findings point to the possibility that the genetic architectures of mental health and physical activity may overlap. The degree and phenotypic specificity of overlap remains to be tested.

### 1.3 Shared brain and genetic architectures?

Together, the studies reviewed above show that mental health and physical activity both involve large-scale brain networks such as the DMN, SN and CEN. Additionally, mental health and physical activity may have partially overlapping genetic architectures, with evidence showing associations between genetic liabilities of psychiatric disorders and physical activity, as well as genetic associations between latent factors of psychopathology and physical activity. As mental health and physical activity are also tightly related at the behavioral level (e.g., emotion, cognition), it is reasonable to speculate that these two constructs may partially overlap in the underlying neurobiological mechanisms. In this study, we aim to determine whether mental health and physical activity have shared variance in brain and genetic architectures, using brain network measures and genomic summary statistics.

## 2. Methods and Materials

### 2.1 Participants

The UK Biobank is an openly accessible population dataset with neuroimaging data collection, in addition to extensive demographic, behavioral, lifestyle, and cognitive measures^8,46^. An initial sample of N=8,378 participants from the UK Biobank (UKB) was considered for this study. These participants had participated in accelerometer-based physical activity evaluations and visited the assessment center, where the resting-state fMRI and mental health data were acquired. Data quality assurance resulted in exclusion of N=64 participants for insufficient accelerometer data (see details below in section 2.2.2), and N=1,925 participants with considerable missing data in the mental health questionnaire (see details below in section 2.2.1). The final sample had N=6,389 participants with 2,994 (46.9%) females (sample mean age=63.74 ± SD 7.53). All participants provided informed consent. UK Biobank has ethical approval from the North West Multi-Centre Research Ethics Committee (MREC). Data access was obtained under UK Biobank application ID 47267.

### 2.2 Data Acquisition and Preprocessing

#### 2.2.1 General mental health measures

The UKB general mental health questionnaire consisting of 41 items was conducted on the same day as fMRI data acquisition (https://biobank.ndph.ox.ac.uk/showcase/label.cgi?id=100060). This self-reported questionnaire primarily measured depression, anxiety, and neuroticism, as well as subjective well-being. These variables had varying degrees of missing data points partly due to responses such as “do not know”, “prefer not to answer”, or “none of above,” and partly due to question dependencies. To ensure robust model estimation while maximizing statistical power, individual variables or questions that had more than 30% missing values were excluded (N=10; see full descriptions for each included individual question in *Table S1*). A multivariate imputation procedure was then leveraged to handle the missingness in the remaining data. By default, this procedure implements multiple imputations with separate imputation models for each incomplete variable^47^. Predictive mean matching (PMM) approach was employed for imputing continuous variables, which first estimates a linear regression model for the target variable (e.g., Y) from all other variables in the data (e.g., non-Y variables) with complete observations. New coefficients are then drawn from the posterior predictive distribution of the estimated regression coefficients and used to calculate the predicted values for the missing entries in Y. The predictive values for the observed Y are also calculated using the estimated regression coefficients. Finally, a small set of candidate donors is formed from the observed Y (i.e., usually 3 or 5 donors) that have the closest predicted values to the missing Y, and the observed value from one donor will be randomly selected to replace the missing value^47^. Using PMM, 20 iterations was performed for each incomplete variable of mental health, and the final imputed value for any given missing entry was averaged across all iterations.

As the current mental health questionnaire covers a broad range of multiple constructs, including depression and anxiety symptoms, neuroticism, and subjective well-being, we performed data decomposition using principal component analysis (PCA) on the imputed data to extract the most relevant information about general psychopathology, using a R package (see details below in section 2.4). We retained the top principal components that collectively explained more than 50% variance of the data in the subsequent statistical analyses. PCA loadings of each individual question per component can be found in *Table 1*.

**Table 1.** Correlations between mental health and physical activity measures.

| Physical Activity |  | Mental Health |  |  |  |  |  |  |
| --- | --- | --- | --- | --- | --- | --- | --- | --- |
|  |  | PC1 | PC2 | PC3 | PC4 | PC5 | PC6 | PC7 |
| Overall | Sleep Duration | -0.08* | 0.04* | 0.01 | 0.00 | 0.04* | -0.01 | 0.03 |
|  | Sedentary | -0.03* | 0.07* | 0.07* | -0.02 | -0.02 | -0.03 | 0.03* |
|  | Light Tasks | 0.02 | -0.02 | 0.01 | -0.01 | -0.04* | -0.01 | 0.00 |
|  | Moderate | -0.02 | -0.06* | 0.02 | 0.01 | 0.03* | 0.04* | 0.01 |
|  | Walking | 0.01 | -0.01 | 0.02 | -0.03* | -0.06* | 0.00 | 0.04* |
|  | Overall Activity | -0.06* | 0.04* | 0.03* | -0.02 | 0.04* | 0.01 | 0.08* |
|  | MET | -0.06* | 0.10* | 0.07* | -0.03 | -0.01 | -0.05* | 0.02 |
| Weekdays | Sleep Duration | -0.08* | 0.04* | 0.02 | 0.00 | 0.03* | -0.01 | 0.03* |
|  | Sedentary | -0.05* | 0.08* | 0.08* | -0.01 | -0.04* | -0.02 | 0.04* |
|  | Light Tasks | 0.01 | -0.02 | 0.03 | -0.03 | -0.03* | -0.01 | 0.01 |
|  | Moderate | -0.02 | -0.05* | 0.03 | 0.01 | 0.02 | 0.03* | 0.03 |
|  | Walking | 0.02 | 0.00 | 0.04* | -0.04* | -0.09* | -0.01 | 0.04* |
|  | Overall Activity | -0.05* | 0.04* | 0.06* | -0.04* | -0.01 | 0.00 | 0.07* |
|  | MET | -0.06* | 0.11* | 0.08* | -0.03 | -0.07* | -0.04* | 0.02 |
| Weekend | Sleep Duration | -0.06* | 0.04* | 0.00 | 0.00 | 0.02 | -0.01 | 0.02 |
|  | Sedentary | -0.02 | 0.05* | 0.06* | -0.02 | -0.01 | -0.02 | 0.02 |
|  | Light Tasks | 0.02 | -0.01 | 0.00 | -0.01 | -0.05* | -0.02 | -0.02 |
|  | Moderate | -0.02 | -0.04* | 0.01 | 0.00 | 0.01 | 0.04* | 0.00 |
|  | Walking | 0.02 | 0.00 | 0.02 | -0.03* | -0.06* | 0.00 | 0.04* |
|  | Overall Activity | -0.05* | 0.04* | 0.03 | -0.02 | 0.03* | 0.01 | 0.06* |
|  | MET | -0.05* | 0.08* | 0.05* | -0.02 | 0.01 | -0.04* | 0.02 |

Phenotypes of mental health were represented by the first seven principal components (i.e., PC1-PC7) that altogether explained 51.05% variance. Significant correlations are indicated by \* after FDR corrections (i.e., all corrected $p$ 's $\leq 0.045$ ) and negative correlation coefficients are highlighted with light grey shading

#### 2.2.2 Physical activity measures

Accelerometer data were acquired for a subset of UKB participants during a seven-day monitoring period (https://biobank.ndph.ox.ac.uk/ukb/label.cgi?id=1008). This enabled real-time measuring of physical activity for the participants throughout the entire week. Following a recommendation for quality control ^48^, data from participants who had less than 72h device wearing time or had no data in each one-hour period of the 24h cycle were excluded (N=64). Using a publicly available machine learning algorithm, we extracted measures of five types of physical activity including sleep, sedentary, walking, light task, and moderate activities^48^. This algorithm applied random forest and hidden Markov models to a 126-dimental vector that represented a range of time and frequency domain features for every non-overlapping 30-sec epoch. The resulting probability of each physical activity was then defined as the count of predicted activity type per 30-sec epoch divvied by the number of epochs^48^. In addition to these probability measures, the average acceleration magnitude, and metabolic equivalents of task (MET) were included to indicate overall activity intensity. The mean values of these features were calculated across weekdays and weekends, respectively, as well as across the entire monitoring period (i.e., average over weekdays and weekends). To account for variation in each individual physical activity measure at different time points, all mean values were standardized by standard deviations for each participant. In total, 21 standardized physical activity measures were included in our analyses.

#### 2.2.3 Resting-state fMRI data and brain network measure

Resting-state fMRI was acquired using a multiband sequence with an acceleration factor of 8 (TR=0.735; voxel size=2.4×2.4×2.4mm3). Preprocessing steps included motion correction, grand-mean intensity normalization, high-pass temporal filtering, unwarping and ICA-FIX denoising (Alfaro-Almagro et al., 2018). Full details can be found in UK Biobank Brain Imaging Documentation (https://biobank.ctsu.ox.ac.uk/crystal/crystal/docs/brain_mri.pdf).

For this study, we used IDPs (imaging-derived phenotypes) that were generated and released by the UKB ^46^. Specifically, partial connectivity matrices and network amplitudes from ICA with dimensionalities of 100 (ICA100) were considered (see *Supplementary Results* for the comparison with ICA25). ICA is a data-driven approach that can estimate the resting-state brain networks reliably and reproducibly, including the three networks of interest. It further helps eliminate noise in the data by separating noise and signal components ^49^. These advantages of ICA make it an unbiased and powerful technique to study the resting-state networks. The mapping of the DMN, SN and CEN onto the ICA components and the calculation of partial connectivity matrices and network amplitudes are described below.

##### 2.2.3.1 Mapping the networks of interest

We used the Stanford FIND atlas ^50^ to construct canonical spatial maps as the reference to identify the three brain networks of interest, namely the DMN, SN and CEN in our sample. Notably, the FIND atlas is a functional connectivity-based parcellation atlas, from which we selected seven functional parcels that are all part of these three intrinsic, including dorsal (i.e. posterior cingulate cortex and medial prefrontal cortex) and ventral default mode networks (i.e., retrosplenial cortex and medial temporal lobe), precuneus network, anterior (i.e., anterior insula and dorsal anterior cingulate cortex) and posterior salience networks (i.e., posterior insula), as well as left and right executive control networks (i.e., dorsolateral prefrontal cortex and parietal cortex in the left and right hemispheres respectively). These selected parcels well represent the subsystems of the three intrinsic networks that have been associated with physical exercises (e.g., via cognitive and interoceptive processing) ^5,6^ and various psychiatric disorders including depression and anxiety ^7–9^. We therefore selected them as our reference networks. To identify ICA components that can be mapped onto these networks of interest, we examined fifty-five signal components that were generated from ICA100 and detected the best matching components corresponding to each of seven selected network parcels, based on spatial correlations. Seven ICA components showing the highest spatial correlations with the FIND atlas were identified (mean r = 0.37). We refer to these ICA components as ‘subnetworks’ from the large-scale DMN, SN and CEN as they represented the subsystems of the three intrinsic networks.

##### 2.2.3.2 Extracting subnetwork edges

Dual regression was performed to obtain timeseries for each ICA component or subnetwork at the individual level ^56^. These extracted subnetwork timeseries were then used to calculate the partial connectivity matrix and amplitude measures of interest. As described in Miller et al.^46^, pairwise partial correlation coefficient were estimated using L2-regularized partial correlations between all 55 signal components from ICA100, which are deemed to be non-artifactual. This analysis resulted in a 55×55 partial correlation matrix, from which we selected the correlation coefficients corresponding to our seven components of interest (i.e., subnetwork edges). In total, 21 subnetwork edges were extracted and included in the subsequent analyses.

##### 2.2.3.3 Calculating subnetwork amplitudes

In additional to pairwise partial correlations between these 7 subnetworks, we further considered the signal amplitude of each subnetwork as the brain network variables. The amplitude is defined as the standard deviation of the ICA component timeseries ^36^. Previous work has shown that amplitudes capture the overall signal fluctuations within each subnetwork and can offer complementary information in relation to behavioral measures. For example, a recent study using the UK Biobank dataset demonstrated independent associations between network amplitudes and behavioral measures in addition to connectivity strength ^46^. Thus, we also included seven subnetwork amplitudes in the subsequent analyses.

##### 2.2.3.4 Overview of resting state measures

In summary, subnetwork edges from the partial connectivity matrix indicate the connection strength between each pair of 7 subnetworks while controlling for all other ICA components, whereas subnetwork amplitudes capture the variance of signal changes within each subnetwork. In total, 28 brain measures including 21 partial connectivity strength measures (i.e., subnetwork edges) and 7 amplitudes were included (see the overlayed subnetworks in *Figure 1;* also see subnetwork selection in *Figure S1*). All of these resting-state imaging measures are available from the UK Biobank showcase (bulk field IDs 25753 and 25755).

### 2.3 Statistical Analysis

Separate statistical analyses were performed to examine the shared brain network architecture and genetic architecture between mental health and physical activity (see *Figure 1* for an overview for the relevant variables and analyses).

#### 2.3.1 Shared variance between mental health and physical activity

Pearson’s correlation was used to identify the shared variance between phenotypes of mental health and physical activity. As we decomposed the data of mental health into principal components, all correlations were performed using the individual-specific component scores. We further calculated the false discovery rate (FDR) to account for multiple testing on all pairwise correlations between mental health and physical activity phenotypes.

#### 2.3.2 Brain Associations with mental health and physical activity

Canonical Correlation Analysis (CCA) has been recognized as a key tool for population neuroimaging that allows for investigating associations between imaging and non-imaging variables^57^. Here in this study, CCA was performed to investigate the associations of brain measures with physical activity and with mental health separately (i.e., simple CCA models). Specifically, CCA finds a linear combination of brain measures that is maximally correlated with a linear combination of mental health or physical activity variables respectively, as defined in *Y* * *A* = *U* ∼ *V* = *X* * *B*^58^ where *Y* is the set of brain measures, *X* the set of mental health or physical activity measures, *A* and *B* are the linear weights, and *U* and *V* the canonical variables or canonical variate pair. The canonical correlation for each pair of canonical variates is defined as the correlation between *U* and *V*. Canonical loadings that indicate the shared variance between the original observations and canonical variables are calculated as the correlations between *Y* and *U*, or between *X* and *V*.

To further identify unique brain associations with physical activity and with mental health respectively, variance in brain network data explained by one set of variables was partialled out in the CCA model for the other set of variables (i.e., the unique CCA model included physical activity measures as covariates in the model for assessing brain-mental health associations and vice versa). Confounding variables were included in all four CCA models and statistical inference for CCA results was made via 1,000 permutations (i.e., breaking correspondence of participant identity with brain measures and mental health/physical activity measures), as implemented in the permCCA package^57^. Notably, the CCA model with the largest number of non-confounding variables included 28 brain measures and 21 physical activity measures, resulting in a ratio of approximately 130 observations (i.e., individuals) per feature. This is expected to ensure sufficient stability for our study^59^.

To further investigate whether the patterns of brain measures in relation to mental health and physical activity overlap, post-hoc analyses were carried out to test the significance of canonical loadings for each individual brain variable. Specifically, we aimed to determine whether the same brain measures contributed significantly to the canonical associations both with mental health and physical activity, and thus could indicate a shared brain basis. These analyses were conducted only for significant canonical variates within each individual CCA model using permutation testing, where correspondence between brain measures and mental health/physical activity measures for each individual participant is shuffled. Canonical loading for each brain variable was recorded per permutation, which resulted in separate null distributions of loadings for each brain variable. The loadings from the true (unpermuted) CCA were then compared against the matching null distributions for each individual brain variable. Statistical significance was determined as the proportion of permuted loadings equal or higher than the observed loadings from the unpermuted analysis, divided by the total number of 1,000 permutations. These permutation-derived p values were further corrected for the number of significant canonical variates within each model (i.e., record the permuted loadings across canonical variates). To compare brain variable patterns across different CCA models, we matched the first significant canonical variates from each model based on the correlations between the canonical variate for the brain measures (i.e., correlating the vector *U* obtained from the mental health CCA models with the vector *U* obtained from the physical activity CCA models).

To characterize the individual mental health questions and physical activity types in relation to the tested brain associations, we further examined the loading patterns of each individual question and physical activity type for the first canonical variate from all models, without testing for statistical significance.

#### 2.3.3 Shared genetic architecture between mental health and physical activity

Genetic correlations between mental health and physical activity were examined by leveraging GWAS summary statistics for the relevant phenotypes.

##### 2.3.3.1 Summary statistics

The mental health questionnaire used in this study includes items that measure neuroticism, anxiety, subjective well-being, depression, and risk taking. We therefore sought to obtain summary statistics for these psychopathological phenotypes. First, we extracted summary statistics for *Neuroticism* from a GWAS meta-analysis of self-reported neuroticism in the UKB (using the same questions as in our study) and Psychiatric Genetics Consortium (using the NEO-FFI personality inventory)^60^. For *Generalized Anxiety Disorder*, we leveraged summary statistics from a GWAS of self-reported Generalized Anxiety Disorder 2-item scale scores in the Million Veteran Program^61^. We further obtained summary statistics for *Subjective Well-Being* from a GWAS meta-analysis of life satisfaction, positive affect, or both life satisfaction and positive affect across 59 cohorts^62^. For *Major Depressive Disorder*, we meta-analyzed summary statistics from case-control GWAS in the UK Biobank and Psychiatric Genomics Consortium^63^ and the Million Veteran Program^64^ (see *Supplementary Results* for further details). Lastly, we obtained summary statistics for *Risk Taking* from a GWAS study using the UKB data, which included the same question of risk taking as in our study^65^. Although these psychopathological phenotypes obtained from the independent GWAS studies were not directly equivalent to the phenotypes derived from mental health questionnaire in our brain association analyses due to different measurements, the underlying constructs of depression, anxiety disorders, and neuroticism, subjective well-being and risk-taking are identical. The phenotypes obtained to conduct genetic association analyses are therefore similar to those in the brain association analyses.

Summary statistics for accelerometer data-derived physical activity phenotypes, including moderate activity, overall activity, sedentary activity, walking, and sleep duration were derived from a GWAS of N=91,105 participants of European ancestry in the UK Biobank^48^. The exact phenotypes were also included in our brain association analyses.

Please refer to *Table 2* for an overview for all the summary statistics used in this study, including sample size and SNP heritability.

**Table 2.** Summary statistics from GWAS studies.

| Reference | Phenotype | Dataset | Sample Size | SNP $h^2$ |
| --- | --- | --- | --- | --- |
| Nagel et al 2018<sup>60</sup> | Neuroticism | UK Biobank & PGC | 390,279 | 0.10 |
| Levey et al 2020<sup>61</sup> | GAD | Million Veteran Program | 199,611 | 0.056 |
| Okbay et al 2016<sup>62</sup> | SWB | Meta-analysis across 59 cohorts | 298,420 | 0.040 |
| Howard et al 2019<sup>63</sup> | MDD | UK Biobank & PGC | 500,199 | 0.11 |
| Levey et al 2021<sup>64</sup> |  | Million Veteran Program | 250,215 |  |
| Linnér et al 2019<sup>65</sup> | Risk-taking | UK Biobank | 431,126 | 0.050 |
| Doherty et al 2018<sup>48</sup> | Physical activities | UK Biobank | 91,105 | 0.10 – 0.21 |

GAD = general anxiety disorder; SWB = subjective well-being; MDD = major depressive disorder; PGC = Psychiatric Genetics Consortium.

##### 2.3.3.2 Genetic correlations

We used linkage disequilibrium score regression (LDSR) and genomic structural equation modeling (gSEM) to test whether the genomic architecture associated with general mental health is shared with physical activity. LDSR leverages GWAS summary statistics to estimate genetic correlations by regressing the SNP statistics on the SNP linkage disequilibrium (LD) scores, or correlations between nearby genomic loci due to population stratification (i.e., systematic differences in allele frequencies due to differences in ancestry). gSEM characterizes the latent genetic architecture across phenotypes based on the LDSR-derived genetic correlation matrices ^66^. To this end, we first applied LDSR to existing GWAS summary statistics of psychopathological phenotypes (i.e., neuroticism, generalized anxiety disorder, subjective well-being, major depressive disorder, and risk taking) and physical activity phenotypes (i.e., overall activity, moderate activity, sedentary activity, sleep duration, and walking), respectively, to estimate pairwise genetic correlations within each construct (i.e., within mental health and within physical activity respectively). We also examined genetic correlations between mental health phenotypes and physical activity phenotypes adjusted for sex and BMI. We then applied gSEM to the covariance matrix of psychopathology and that of physical activity separately, allowing one single latent factor to load freely within each model. Metrics indicating model fit (i.e., CFI, comparative fit index; SRMR, standardized root mean squared residual) and factor loadings from each of these models were used to determine whether one common genetic factor fit the physical activity and mental health data well, respectively.

Because our results indicated poor model fit for some gSEM analyses (see details below in Results section 3.3), we focused on the model of mental health, where “risk-taking” was excluded to generate a latent factor of “negative affect” across other phenotypes. Specifically, we explored genetic correlations between the latent factor of “negative affect” (i.e., without risk-taking) and each of the five physical activity phenotypes. In addition, we examined genetic correlations between “risk-taking” alone and each individual physical activity phenotype, using LDSR. False discovery rate correction was used to correct for multiple testing (N=10 tests). We also repeated analyses that returned significant results, with adjustment for sex and BMI. Adjusted summary statistics from Doherty and colleagues (2018) were used in these analyses.

##### 2.3.3.3 Causal relationships

To examine plausible causal associations between physical activity phenotypes and negative affect, we conducted Latent Causal Variable Analysis (LCV). This approach finds a latent variable that mediates the genetic correlation between two traits, such as negative affect and sleep duration. Generally, if the latent variable has a stronger genetic correlation with trait 1 (e.g., sleep duration) than with trait 2 (e.g., negative affect), part of the genetic component of trait 1 is thought to be causal for trait 2. This partial causality can be quantified using the genetic causality proportion (GCP) of trait 1 (sleep duration) on trait 2 (negative affect), which can range between 0 (no partial genetic causality) and 1 (full genetic causality) ^67^. The advantages of using LCV over the more traditional Mendelian Randomization for causal inference include increased power by leveraging SNPs across the genome and less susceptibility to confounding by horizontal pleiotropy ^67^.

### 2.4 Confounding variables

Based on the literature, BMI, smoking and drinking status were included as confounding factors in all statistical analyses for brain associations^68,69^. Age, sex, head motion during rs-fMRI acquisition (i.e., mean frame-wise displacement), time difference in days between accelerometer recording (i.e., start date) and assessment center visit date (i.e., acquisition date for both mental health questionnaire and rs-fMRI), as well as the scanning site were further included. Due to varying degrees of missingness in the confound variables (i.e., up to 22%), the same imputation procedure, as described in *Section 2.2.2*, was performed except that additional usage of multinomial logistic regression was employed to impute categorical data with more than two levels (i.e., smoking and drinking status). Complete observations from all variables including the confounding variables were used in the imputation procedure. When calculating the final predicted values to replace missing data, the predicted values for continuous variables were averaged across 20 iterations and the level with the highest count across iterations was selected. All categorical variables after imputation were dummy coded for subsequent statistical analyses. In the genetic correlation analyses, to ensure robust test effects, we repeated models with significant results including sex and BMI as covariates.

### 2.5 Data and Code Availability

The UK Biobank data used in this study can be accessed by researchers upon application (https://www.ukbiobank.ac.uk/register-apply). A machine learning algorithm shared on github was used to extract physical activity measures (https://github.com/activityMonitoring/biobankAccelerometerAnalysis). The Matlab code for running permutation inference for CCA are also available on github, including the new extension for testing canonical variable loadings (https://github.com/andersonwinkler/PermCCA). We performed Pearson’s correlations and imputations in R (version 4.1.0) ^70^, using basic *stat* function and *MICE* package ^47^, respectively. Genetic analyses including LDSR and gSEM were performed using the GenomicSEM package also in R ^66^, with code available on github (https://github.com/GenomicSEM/GenomicSEM). LCV was performed using the open-source LCV software (https://github.com/lukejoconnor/LCV). We also shared the derived data from statistical analyses and code for producing the figures on the Open Science Framework (https://osf.io/p3fzv/). To note, the GWAS summary statistics used in the current study are hosted elsewhere, for which we only shared the downloading links.

## 3. Results (1,054 words)

### 3.1 Correlations between mental health and physical activity measures

In this sample, phenotypic measures of mental health and physical activity demonstrated overall small but significant correlations, with coefficients ranging between −0.08 and 0.11 (FDR-corrected p-values ≤ 0.045). As shown in *Table 1*, the mental health measures (i.e., principal component scores of the seven principal components) were broadly associated with all physical activity measures. In particular, sleep showed the strongest association with the first principal component (PC1) of mental health such that greater sleep duration was associated with poorer overall mental health (i.e., loadings of PC1 were mostly negative as shown in Table S1) and walking had the greatest association with the fifth principal component (PC5) of mental health such that higher depression scores was linked to reduced walking time. The overall coefficient patterns were largely consistent for correlations between physical activity and mental health measures despite of minor differences across different time windows (i.e., overall, weekdays, weekend; see full correlations in *Table 1*).

### 3.2 Shared neural correlates of mental health and physical activity

#### 3.2.1 Brain associations with mental health and physical activity

In simple CCA models (i.e., without accounting for the potentially shared variance in brain measures between physical activity and mental health), 2 and 3 significant canonical variate pairs were observed respectively for brain-mental health (r_1_=0.16, p_1_=0.001; r_2_=0.12, p_2_=0.001) and brain-physical activity associations (r_1_=0.23, p_1_=0.001; r_2_=0.15, p_2_=0.001; r_3_=0.13, p_3_=0.047). When controlling for the shared variance in brain measures between mental health and physical activity (i.e., unique models), we found 2 significant canonical variates for both brain-mental health (r_1_=0.14, p_1_=0.001; r_2_=0.12, p_2_=0.002) and brain-physical activity associations (r_1_=0.21, p_1_=0.001; r_2_=0.15, p_2_=0.001), with slightly decreased canonical correlation coefficients comparing to those from the simple models. The canonical variates for brain measures after accounting for the shared variance (i.e., from the unique models) mapped well with those from the simple models, as indicated by correlation coefficients between brain canonical variables (i.e., □; all r’s > 0.96; see full results in *Table S2*). These results are in line with our expectations that both mental health and physical activity are closely associated with the functional networks under investigation and that mental health and physical activity have shared variance in the functioning of these networks, as indicated by the reduced canonical correlation coefficients from the unique models.

#### 3.2.2 Canonical loadings of brain measures

Findings from the post-hoc analyses on the significant canonical variates showed considerable overlap between the brain measures with significant loadings associated with mental health and with physical activity. Overall, the amplitude of subnetworks that indicates the overall signal fluctuations in each subnetwork over time loaded higher than the edges, with the highest loadings on the amplitude of dorsal DMN or left CEN for the first canonical variates in both the mental health and physical activity models (*permuted p-values* ≤ 0.001; *Figure 2*). This suggested that the magnitude of fluctuations of intra-network signal (i.e., amplitude) had higher contributions to the observed canonical associations with both mental health and physical activity than the inter-network connectivity strength. The similar loading pattern of all brain measures were largely retained even when the shared variance was partialled out for mental health and for physical activity, respectively, in the unique models (*permuted* p-values ≤ 0.035). In addition to amplitude, connectivity between the dorsal DMN and left CEN (i.e., subnetwork edge) also exhibited statistically significant loadings in the first canonical variates of both the mental health and the physical activity models (simple and unique models; *permuted* p-values ≤ 0.001; *Figure 2*). The brain variable loadings for the second canonical variate were highly similar between simple and unique models for either the mental health or physical activity models, with the amplitude and subnetwork edges showing evenly important involvement. Yet, the patterns of these loadings differed between the mental health and physical activity model (*Figure S2*).

#### 3.2.3 Canonical loadings of mental health questions and physical activities

Canonical loadings of both individual mental health questions and physical activity types for the first canonical variate also exhibited similar patterns between the simple and unique models (*Figure S3*). In the models for brain-mental health associations, “risk taking” and “ever irritable/ argumentative for 2 days” had the highest loadings in both simple and unique models, whereas in the models for brain-physical activity associations, “walking” in all time windows (i.e., overall, weekdays, weekend) showed the greatest importance across models. Interestingly, among all physical activity types, only “sleep” showed the opposite direction in canonical loadings (*Figure S3*).

### 3.3 Genetic correlations

#### 3.3.1 Genetic correlations for individual phenotypes

Pairwise LDSR was performed separately for psychopathological phenotypes and physical activity phenotypes. All results were significant after controlling for multiple comparisons, except for the genetic correlations between neuroticism and risk-taking (*rg* = 0.039, *p* = 0.075), and between subjective well-being and risk-taking (*rg* = 0.054, *p* = 0.146; *Figure 3*). Specifically, among psychopathological phenotypes, the smallest significant genetic correlation was observed for generalized anxiety disorder and risk-taking (*rg* = 0.151, *p* = 2.69e-05, *FDR-corrected p-value =* 3.36e-05), and the largest effect was observed for MDD and generalized anxiety disorder (*rg* = 0.768, *p* = 2.62e-90, *FDR-corrected p-value =* 1.31e-89). For physical activity, genetic correlations ranged from -0.217 (*p* = 1.03e-04, *FDR-corrected p-value* = 1.03e-04) for sleep duration and walking to 0.796 (*p* = 1.77e-31, *FDR-corrected p-value* = 8.85e-31) for moderate and overall activity (*Figure 3*). Physical activity phenotypes adjusted for sex and BMI largely recapitulated these results, with genetic correlations ranging from -0.209 (*p* = 2.49e-04, *FDR-corrected p-value =* 2.49e-04) for sleep duration and walking, to 0.780 (*p* = 2.12e-23, *FDR-corrected p-value* = 1.06e-22) for moderate and overall activities.

#### 3.3.2 Genetic correlations for latent factors

Analyses using gSEM across all mental health phenotypes returned results indicating suboptimal estimations with a low factor loading on risk taking from the model for a general psychopathology latent factor (standardized loading = 0.065), which likely reflected its conceptually distinct construct from all other phenotypes (i.e., depression, anxiety, neuroticism, and subjective well-being). Additionally, initial gSEM results for physical activity phenotypes indicated overall poor model fit (CFI = 0.569, SRMR = 0.160; *Figure S4*). Thus, we performed gSEM for psychopathological phenotypes grouped as one “negative affect” latent factor after excluding “risk-taking”, with an effective sample size of 571,170 and good model fit (CFI = 0.987, SRMR = 0.0535; *Figure S5*). After correction for multiple testing, genetic variance in negative affect was significantly and positively correlated with sleep duration (*rg* = 0.121, *p* = 1.25e-05, *FDR-corrected p* = 1.25e-04), and negatively correlated with moderate (*rg* = -0.117, *p* = 1.33e-03, *FDR-corrected p* = 4.43e-03) and overall activity (*rg* = -0.085, *p* = 7.70e-04, *FDR-corrected p* = 3.85e-03; *Figure 4;* also see the results for a latent negative affect factor including “risk-taking” in *the Supplementary Results*). No significant genetic correlation was observed between negative affect and sedentary activity or walking after multiple comparison corrections (|rgs| ≤ 0.062, p-values ≥ 0.036, FDR-corrected p-values ≥ 0.060; *Table 3*). Post-hoc analyses revealed that after adjustment for sex and BMI, only the genetic correlation between negative affect and sleep duration remained significant (*rg* = 0.122, *p* = 9.38e-06). Using LDSR, risk-taking was genetically correlated with both overall activity and walking (|rgs| ≥ 0.074, *p*s ≤ 0.020, *ps*FDR ≤ 0.040) but not moderate or sedentary activity or sleep duration (|rgs| ≤ 0.081, *p*s ≥ 0.092).

**Table 3.** Genetic correlations between mental health and physical activity phenotypes.

| Mental Health Phenotype | Physical Activity Phenotype | *rg* | *p* | *pFDR* |
| --- | --- | --- | --- | --- |
| Risk Tolerance | Sleep Duration | -0.029 | 0.433 | 0.447 |
|  | Sedentary | -0.028 | 0.447 | 0.447 |
|  | Moderate | 0.081 | 0.092 | 0.131 |
|  | Walking | **0.115** | 4.95e-3 | 0.012 |
|  | Overall | **0.074** | 0.020 | 0.040 |
| Negative Affect | Sleep Duration | **0.121** | 1.25e-5 | 1.25e-4 |
|  | Sedentary | -0.062 | 0.036 | 0.060 |
|  | Moderate | **-0.117** | 1.33e-3 | 4.43e-3 |
|  | Walking | -0.029 | 0.383 | 0.447 |
|  | Overall | **-0.085** | 7.70e-4 | 3.85e-3 |

*Note*, Significant results after FDR corrections were highlighted in bold. *rg* = genetic correlation, *pFDR* = FDR-corrected p-value.

#### 3.3.3 Latent Causal Variable Analysis

Results from the LCV analyses did not indicate causal relationships between any physical activity phenotypes and negative affect, in either direction (|GCPs| < 0.425; ps > 0.163).

## 4. Discussion (1,107 words)

In this study, we investigated whether mental health and physical activity have shared brain and genetic architectures using the UK Biobank cohort. Our findings showed significant associations of mental health and physical activity separately with a set of brain measures that represent the connectivity strength and amplitude of subnetworks from the DMN, SN and CEN. Critically, these significant associations exhibited highly similar patterns of brain variable loadings across mental health and physical activity models even when the shared variance between these two constructs was accounted for, suggesting a potential overlap in brain network architecture between these two constructs. Further analyses examining genetic correlations for mental health and physical activity showed that negative affect exhibited significant genetic correlations with several physical activity types, of which sleep duration demonstrated the strongest genetic correlation that remained significant after controlling for BMI and sex effects. Together, these results support the presence of shared multivariate brain and genetic architectures between mental health and physical activity.

The three intrinsic brain networks, namely the DMN, SN and CEN, have been consistently implicated in a wide range of psychiatric disorders including major depression and anxiety^10^. Interestingly, connections between or the configurations of these networks have also been associated with physical exercises^71,72^. The current study therefore focused specifically on the subnetworks from these large-scale networks and used the amplitude and connectivity strength (i.e., subnetwork edge) to examine the associations of these networks with mental health and with physical activity, respectively. In line with the literature, we observed significant multivariate associations for all three networks with either mental health or physical activity, and significant loadings on most of the subnetworks. In particular, the dorsal DMN and left CEN showed the greatest involvement in the observed brain associations with both mental health and physical activity (*Figure 2*). In this study, the dorsal DMN subnetwork primarily consisted of the posterior cingulate cortex (PCC) and the ventromedial prefrontal cortex (vmPFC), the two brain areas that are commonly considered as the core subsystem of the DMN^73^. Similarly, the subnetwork of the CEN here included two most typical hub regions: the dorsolateral prefrontal cortex (dlPFC) and posterior parietal cortex (PPC) for each individual hemisphere (*Figure S1*). These major hubs of the DMN and CEN have been implicated in various mental illnesses including depression and anxiety. For example, the PCC and mPFC have been suggested to collectively support multiple cognitive functions including decision making and memory^74^, the impairment of which has often been reported in patients with major depression and anxiety disorders^75–77^. Additionally, the dlPFC is known to be involved in emotion regulation ^78–80^ and dysfunction of this region is often seen in abnormal processing of emotional experiences in patients with depressive and anxiety symptoms^81,82^. As for physical activity, increased dlPFC activity has been observed after acute physical exercises in participants with higher scores in the Stroop test^83^, whereas the involvement of the DMN subsystems in the medial temporal lobe (MTL) including the hippocampus and its connection with the medial PFC are often observed in relation to enhanced memory after physical exercises^20,84,85^. In our findings, the amplitude of dorsal DMN and left CEN that indicated the magnitude of fluctuations of intra-network signal (i.e., variance in the connections between PCC and vmPFC or between dlPFC and PCC), as well as the connectivity strength between these two subnetworks, showed significantly high loadings for the most critical association between brain measures and mental health measures (i.e., the first canonical variate). These observations are in line with separate literature on mental health and physical activity, and provide empirical evidence that mental health and physical activity may share brain architecture involved in major cognitive functions.

Interestingly, mental health and physical activity also appear to have partially overlapping genetic architectures. In line with previous reports that internalizing problems are negatively genetically correlated with physical movement^45^, we showed that a latent negative affect factor capturing genetic covariance between subjective well-being, neuroticism, major depressive disorder, and generalized anxiety disorder was negatively genetically correlated with overall physical activity as well as a more fine-grained phenotype of moderate activity, and positively genetically correlated with sleep duration. Protective effects of physical activity on mental health have long been documented, as have negative health consequences of psychiatric disorders^5–7,86^. Here, we demonstrate that these relationships can be partially explained by shared genetic p redisposition, although results from our latent causal variable analysis indicate that these associations do not reflect causal influences. Interestingly, with the adjustment of sex and BMI, only the correlation with sleep duration remained significant. Sleep duration also showed the lowest loading onto a latent physical activity genetic factor (*Figure S4*). These results suggest that the sleep phenotype is somewhat distinct from the remaining physical activity phenotypes and that the shared genetic architecture between negative affect and sleep duration is more pronounced than that between negative affect and the degree of daily physical or sedentary activity. This is in line with the frequent documentation of symptomatic sleep disturbances across forms of psychopathology, including depression and anxiety^87,88^, even in children^89^. Specifically, this aligns well with the literature showing reduced sleep duration in older adults with depression and anxiety disorders ^90–92^

Despite being the first to jointly investigate the shared brain network architecture and genetic basis of mental health and physical activity in a large population cohort, our study has some limitations. First, the brain measures in our study are derived from resting-state fMRI measures. Although our choice reflected a rich literature that has implicated these measures in both mental health and physical activity, addressing smaller-scale brain structures (e.g., specific regions) with relevant hypotheses and inclusion of multimodal brain measures such as structural gray matter volume, cortical thickness, and white matter integrity can provide complementary insights into brain architecture in relation to mental health and physical activity, and thus may be of interest for future investigations. Second, although the accelerometer recording took place prior to the acquisition of resting-state fMRI and mental health assessment for most of the participants in our study (i.e., 96%), the degrees of time difference between these measurements varied greatly at the individual level (i.e., ranging between -473 to 2281 days). This time discrepancy was accounted for in all CCA models as a covariate and cautions should be taken when interpreting the observed brain associations with reference to time effects. It should also be noted that the mean age of the current sample is relatively high as the UKB cohort comprises predominantly middle-to-late aged individuals. Our findings therefore should be interpreted carefully in the relevant context. Lastly, socioeconomic variables including education attainment can be relevant to mental health and physical activity phenotypes and inclusion of these variables as confounding factors may be considered in future investigations.

In conclusion, our study jointly analyzed resting-state network measures and genetic correlations in a large cohort to test the hypothesis of a shared neurobiological basis of mental health and physical activity. Our findings revealed that multivariate patterns of brain correlates were highly similar between mental health and physical activity and highlighted genetic correlations between mental health (negative affect) and overall physical activity, moderate activity levels, and sleep duration. Taken together, these findings point towards neural and genetic mechanisms that may subserve the protective influence of physical exercise and sleep on mental health.

## Data Availability

The UK Biobank data used in this study can be accessed by researchers upon application (https://www.ukbiobank.ac.uk/register-apply).

## Acknowledgements

JDB is supported by the NIH (1 R34 NS118618-01) and the McDonnell Center for Systems Neuroscience. SEP is supported by the NIH (1 F31 AA029934-01).

## Figure Legends

**Figure 1. Overview of variables and analyses.** Separate analyses were conducted for brain (left panel) and genetic associations (right panel). For brain associations, canonical correlation analysis (CCA) was employed for mental health (MH) and physical activity (PA) separately. Simple and unique CCA models only differed in whether the shared variance between MH and PA was accounted for. For genetic associations, GWAS summary statistics for 5 mental health phenotypes (MHP_1_-MHP_5_) and for 5 physical activity phenotypes (PAP_1_-PAP_5_) were leveraged into pairwise linkage disequilibrium score regression (LDSR) analyses in MHP and PAP models separately. Genomic structural equation modeling (gSEM) was also employed to identify genetic associations between a latent factor from MH phenotypes and each of PA phenotype, followed by latent causal variable analysis (LCV) that allows for inferring causal genetic relationships among MH and PA phenotypes.

**Figure 2. Canonical loadings of brain measures on the first canonical variates.** These loadings represent the linear correlation between the original brain measures (Y) and the first canonical variate (U) per model. Color coding was made for brain variable names along the Y axis (i.e., subnetwork edges in gray with “-” between subnetwork names and amplitude in orange), and for the bars representing canonical loadings (i.e., significance in cyan, insignificance in yellow). Simple and unique models differ in whether the model accounted for the shared variance in brain measures between mental health and physical activity. vDMN = ventral default mode network; R/L CEN = right/ left central executive network; PCu = precuneus; a/p SN = anterior/posterior salience network.

**Figure 3. Heatmap of bivariate genetic correlations.** Pairwise genetic correlations were calculated separately for phenotypes of mental health (A) and those of physical activity (B). rg = genetic correlation coefficient, GAD = Generalized Anxiety Disorder, MDD = Major Depressive Disorder, Risk = Risk Tolerance, SWB = Subjective Well-Being.

**Figure 4. Genetic correlations between negative affect and physical activity phenotypes.** Significant genetic correlations with negative affect were observed for moderate (A), overall activities (B), and sleep duration (C). The latent negative affect factor loaded positively on neuroticism, major depression disorder (MDD) and general anxiety disorder (GAD) phenotypes, but negatively on the subjective well-being (SWB) phenotype.

## Reference

1 Zhang Z, Chen W. A Systematic Review of the Relationship Between Physical Activity and Happiness. J. Happiness Stud. 2019. doi:10.1007/s10902-018-9976-0.

2 Fernandes J, Arida RM, Gomez-Pinilla F. Physical exercise as an epigenetic modulator of brain plasticity and cognition. Neurosci. Biobehav. Rev. 2017. doi:10.1016/j.neubiorev.2017.06.012.

3 Kays JL, Hurley RA, Taber KH. The dynamic brain: Neuroplasticity and mental health. J Neuropsychiatry Clin Neurosci 2012. doi:10.1176/appi.neuropsych.12050109.

4 Millan MJ, Agid Y, Brüne M, Bullmore ET, Carter CS, Clayton NS et al. Cognitive dysfunction in psychiatric disorders: Characteristics, causes and the quest for improved therapy. Nat. Rev. Drug Discov. 2012. doi:10.1038/nrd3628.

5 Biddle S. Physical activity and mental health: Evidence is growing. World Psychiatry. 2016. doi:10.1002/wps.20331.

6 Deslandes A, Moraes H, Ferreira C, Veiga H, Silveira H, Mouta R et al. Exercise and mental health: Many reasons to move. Neuropsychobiology. 2009. doi:10.1159/000223730.

7 Heinze K, Cumming J, Dosanjh A, Palin S, Poulton S, Bagshaw AP et al. Neurobiological evidence of longer-term physical activity interventions on mental health outcomes and cognition in young people: A systematic review of randomised controlled trials. Neurosci. Biobehav. Rev. 2021. doi:10.1016/j.neubiorev.2020.10.014.

8 Sudlow C, Gallacher J, Allen N, Beral V, Burton P, Danesh J et al. UK Biobank: An Open Access Resource for Identifying the Causes of a Wide Range of Complex Diseases of Middle and Old Age. PLoS Med 2015. doi:10.1371/journal.pmed.1001779.

9 Woodward ND, Cascio CJ. Resting-state functional connectivity in psychiatric disorders. JAMA Psychiatry 2015; 72: 743–744.

10 Menon V. Large-scale brain networks and psychopathology: A unifying triple network model. Trends Cogn Sci 2011; 15: 483–506.

11 Tozzi L, Zhang X, Chesnut M, Holt-Gosselin B, Ramirez CA, Williams LM. Reduced functional connectivity of default mode network subsystems in depression: Meta-analytic evidence and relationship with trait rumination. NeuroImage Clin 2021. doi:10.1016/j.nicl.2021.102570.

12 Kaiser RH, Andrews-Hanna JR, Wager TD, Pizzagalli DA. Large-Scale Network Dysfunction in Major Depressive Disorder. JAMA Psychiatry 2015. doi:10.1001/jamapsychiatry.2015.0071.

13 Xu J, Van Dam NT, Feng C, Luo Y, Ai H, Gu R et al. Anxious brain networks: A coordinate-based activation likelihood estimation meta-analysis of resting-state functional connectivity studies in anxiety. Neurosci. Biobehav. Rev. 2019. doi:10.1016/j.neubiorev.2018.11.005.

14 Servaas MN, Geerligs L, Renken RJ, Marsman JBC, Ormel J, Riese H et al. Connectomics and neuroticism: An altered functional network organization. Neuropsychopharmacology 2015. doi:10.1038/npp.2014.169.

15 Colcombe SJ, Kramer AF, Erickson KI, Scalf P, McAuley E, Cohen NJ et al. Cardiovascular fitness, cortical plasticity, and aging. Proc Natl Acad Sci U S A 2004. doi:10.1073/pnas.0400266101.

16 Herold F, Aye N, Lehmann N, Taubert M, Müller NG. The contribution of functional magnetic resonance imaging to the understanding of the effects of acute physical exercise on cognition. Brain Sci 2020. doi:10.3390/brainsci10030175.

17 Ikuta T, Loprinzi PD. Association of cardiorespiratory fitness on interhemispheric hippocampal and parahippocampal functional connectivity. Eur J Neurosci 2019. doi:10.1111/ejn.14366.

18 Talukdar T, Nikolaidis A, Zwilling CE, Paul EJ, Hillman CH, Cohen NJ et al. Aerobic fitness explains individual differences in the functional brain connectome of healthy young adults. Cereb Cortex 2018. doi:10.1093/cercor/bhx232.

19 Weng TB, Pierce GL, Darling WG, Falk D, Magnotta VA, Voss MW. The Acute Effects of Aerobic Exercise on the Functional Connectivity of Human Brain Networks. Brain Plast 2016. doi:10.3233/bpl-160039.

20 Voss MW, Prakash RS, Erickson KI, Basak C, Chaddock L, Kim JS et al. Plasticity of brain networks in a randomized intervention trial of exercise training in older adults. Front Aging Neurosci 2010. doi:10.3389/fnagi.2010.00032.

21 Greeley B, Chau B, Jones CB, Neva JL, Kraeutner SN, Campbell KL et al. Multiple bouts of high-intensity interval exercise reverse age-related functional connectivity disruptions without affecting motor learning in older adults. Sci Rep 2021. doi:10.1038/s41598-021-96333-4.

22 Schmitt A, Upadhyay N, Martin JA, Rojas S, Strüder HK, Boecker H. Modulation of Distinct Intrinsic Resting State Brain Networks by Acute Exercise Bouts of Differing Intensity. Brain Plast 2019. doi:10.3233/bpl-190081.

23 Voss MW, Weng TB, Narayana-Kumanan K, Cole RC, Wharff C, Reist L et al. Acute exercise effects predict training change in cognition and connectivity. Med Sci Sports Exerc 2020. doi:10.1249/MSS.0000000000002115.

24 Nutt DJ, Wilson S, Paterson L. Sleep disorders as core symptoms of depression. Dialogues Clin Neurosci 2008. doi:10.31887/dcns.2008.10.3/dnutt.

25 Riemann D, Krone LB, Wulff K, Nissen C. Sleep, insomnia, and depression. Neuropsychopharmacology. 2020. doi:10.1038/s41386-019-0411-y.

26 Cox RC, Olatunji BO. Sleep in the anxiety-related disorders: A meta-analysis of subjective and objective research. Sleep Med. Rev. 2020. doi:10.1016/j.smrv.2020.101282.

27 Mellman TA. Sleep and Anxiety Disorders. Psychiatr. Clin. North Am. 2006. doi:10.1016/j.psc.2006.08.005.

28 Rumble ME, White KH, Benca RM. Sleep Disturbances in Mood Disorders. Psychiatr. Clin. North Am. 2015. doi:10.1016/j.psc.2015.07.006.

29 Chennaoui M, Arnal PJ, Sauvet F, Léger D. Sleep and exercise: A reciprocal issue? Sleep Med. Rev. 2015. doi:10.1016/j.smrv.2014.06.008.

30 Wainberg M, Jones SE, Beaupre LM, Hill SL, Felsky D, Rivas MA et al. Association of accelerometer-derived sleep measures with lifetime psychiatric diagnoses: A cross-sectional study of 89,205 participants from the UK Biobank. PLOS Med 2021; 18: e1003782.

31 Full KM, Malhotra A, Gallo LC, Kerr J, Arredondo EM, Natarajan L et al. Accelerometer-Measured Sleep Duration and Clinical Cardiovascular Risk Factor Scores in Older Women. Journals Gerontol - Ser A Biol Sci Med Sci 2020. doi:10.1093/gerona/glz201.

32 De Havas JA, Parimal S, Soon CS, Chee MWL. Sleep deprivation reduces default mode network connectivity and anti-correlation during rest and task performance. Neuroimage 2012. doi:10.1016/j.neuroimage.2011.08.026.

33 Sämann PG, Tully C, Spoormaker VI, Wetter TC, Holsboer F, Wehrle R et al. Increased sleep pressure reduces resting state functional connectivity. Magn Reson Mater Physics, Biol Med 2010. doi:10.1007/s10334-010-0213-z.

34 Yeo BTT, Tandi J, Chee MWL. Functional connectivity during rested wakefulness predicts vulnerability to sleep deprivation. Neuroimage 2015. doi:10.1016/j.neuroimage.2015.02.018.

35 Khalsa S, Mayhew SD, Przezdzik I, Wilson R, Hale J, Goldstone A et al. Variability in cumulative habitual sleep duration predicts waking functional connectivity. Sleep 2016. doi:10.5665/sleep.5324.

36 Bijsterbosch J, Harrison S, Duff E, Alfaro-Almagro F, Woolrich M, Smith S. Investigations into within-and between-subject resting-state amplitude variations. Neuroimage 2017. doi:10.1016/j.neuroimage.2017.07.014.

37 Anderson E, Shivakumar G. Effects of exercise and physical activity on anxiety. Front Psychiatry 2013. doi:10.3389/fpsyt.2013.00027.

38 Blumenthal JA, Babyak MA, Doraiswamy PM, Watkins L, Hoffman BM, Barbour KA et al. Exercise and pharmacotherapy in the treatment of major depressive disorder. Psychosom Med 2007. doi:10.1097/PSY.0b013e318148c19a.

39 Choi KW, Zheutlin AB, Karlson RA, Wang MJ, Dunn EC, Stein MB et al. Physical activity offsets genetic risk for incident depression assessed via electronic health records in a biobank cohort study. Depress Anxiety 2020. doi:10.1002/da.22967.

40 Dennison CA, Legge SE, Bracher-Smith M, Menzies G, Escott-Price V, Smith DJ et al. Association of genetic liability for psychiatric disorders with accelerometer-assessed physical activity in the UK Biobank. PLoS One 2021. doi:10.1371/journal.pone.0249189.

41 Anttila V, Bulik-Sullivan B, Finucane HK, Walters RK, Bras J, Duncan L et al. Analysis of shared heritability in common disorders of the brain. Science (80-) 2018. doi:10.1126/science.aap8757.

42 Lee SH, Ripke S, Neale BM, Faraone S V., Purcell SM, Perlis RH et al. Genetic relationship between five psychiatric disorders estimated from genome-wide SNPs. Nat Genet 2013. doi:10.1038/ng.2711.

43 Taylor MJ, Martin J, Lu Y, Brikell I, Lundström S, Larsson H et al. Association of Genetic Risk Factors for Psychiatric Disorders and Traits of These Disorders in a Swedish Population Twin Sample. JAMA Psychiatry 2019. doi:10.1001/jamapsychiatry.2018.3652.

44 Smoller JW, Andreassen OA, Edenberg HJ, Faraone S V., Glatt SJ, Kendler KS. Psychiatric genetics and the structure of psychopathology. Mol. Psychiatry. 2019. doi:10.1038/s41380-017-0010-4.

45 Grotzinger AD, Mallard TT, Akingbuwa WA, Ip HF, Adams MJ, Lewis CM et al. Genetic architecture of 11 major psychiatric disorders at biobehavioral, functional genomic, and molecular genetic levels of analysis. medRxiv. 2020. doi:10.1101/2020.09.22.20196089.

46 Miller KL, Alfaro-Almagro F, Bangerter NK, Thomas DL, Yacoub E, Xu J et al. Multimodal population brain imaging in the UK Biobank prospective epidemiological study. Nat Neurosci 2016. doi:10.1038/nn.4393.

47 van Buuren S, Groothuis-Oudshoorn K. mice: Multivariate imputation by chained equations in R. J Stat Softw 2011. doi:10.18637/jss.v045.i03.

48 Doherty A, Smith-Byrne K, Ferreira T, Holmes M V., Holmes C, Pulit SL et al. GWAS identifies 14 loci for device-measured physical activity and sleep duration. Nat Commun 2018; 9. doi:10.1038/s41467-018-07743-4.

49 Griffanti L, Douaud G, Bijsterbosch J, Evangelisti S, Alfaro-Almagro F, Glasser MF et al. Hand classification of fMRI ICA noise components. Neuroimage 2017. doi:10.1016/j.neuroimage.2016.12.036.

50 Shirer WR, Ryali S, Rykhlevskaia E, Menon V, Greicius MD. Decoding Subject-Driven Cognitive States with Whole-Brain Connectivity Patterns. Cereb Cortex 2012; 22: 158–165.

51 Voss MW, Nagamatsu LS, Liu-Ambrose T, Kramer AF. Exercise, brain, and cognition across the life span. J. Appl. Physiol. 2011. doi:10.1152/japplphysiol.00210.2011.

52 Wallman-Jones A, Perakakis P, Tsakiris M, Schmidt M. Physical activity and interoceptive processing: Theoretical considerations for future research. Int. J. Psychophysiol. 2021. doi:10.1016/j.ijpsycho.2021.05.002.

53 Sha Z, Wager TD, Mechelli A, He Y. Common Dysfunction of Large-Scale Neurocognitive Networks Across Psychiatric Disorders. Biol Psychiatry 2019. doi:10.1016/j.biopsych.2018.11.011.

54 Sha Z, Xia M, Lin Q, Cao M, Tang Y, Xu K et al. Meta-connectomic analysis reveals commonly disrupted functional architectures in network modules and connectors across brain disorders. Cereb Cortex 2018. doi:10.1093/cercor/bhx273.

55 Kolesar TA, Bilevicius E, Wilson AD, Kornelsen J. Systematic review and meta-analyses of neural structural and functional differences in generalized anxiety disorder and healthy controls using magnetic resonance imaging. NeuroImage Clin. 2019. doi:10.1016/j.nicl.2019.102016.

56 Nickerson LD, Smith SM, Öngür D, Beckmann CF. Using dual regression to investigate network shape and amplitude in functional connectivity analyses. Front Neurosci 2017; 11. doi:10.3389/fnins.2017.00115.

57 Winkler AM, Renaud O, Smith SM, Nichols TE. Permutation inference for canonical correlation analysis. Neuroimage 2020. doi:10.1016/j.neuroimage.2020.117065.

58 Hotelling H. RELATIONS BETWEEN TWO SETS OF VARIATES. Biometrika 1936. doi:10.1093/biomet/28.3-4.321.

59 Helmer M, Warrington S, Mohammadi-Nejad A-R, Ji LJ, Howell A, Rosand B et al. On stability of Canonical Correlation Analysis and Partial Least Squares with application to brain-behavior associations. bioRxiv 2020.

60 Nagel M, Jansen PR, Stringer S, Watanabe K, De Leeuw CA, Bryois J et al. Meta-analysis of genome-wide association studies for neuroticism in 449,484 individuals identifies novel genetic loci and pathways. Nat Genet 2018. doi:10.1038/s41588-018-0151-7.

61 Levey DF, Gelernter J, Polimanti R, Zhou H, Cheng Z, Aslan M et al. Reproducible Genetic Risk Loci for Anxiety: Results From ∼200,000 Participants in the Million Veteran Program. Am J Psychiatry 2020. doi:10.1176/appi.ajp.2019.19030256.

62 Okbay A, Baselmans BML, De Neve JE, Turley P, Nivard MG, Fontana MA et al. Genetic variants associated with subjective well-being, depressive symptoms, and neuroticism identified through genome-wide analyses. Nat Genet 2016. doi:10.1038/ng.3552.

63 Howard DM, Adams MJ, Clarke TK, Hafferty JD, Gibson J, Shirali M et al. Genome-wide meta-analysis of depression identifies 102 independent variants and highlights the importance of the prefrontal brain regions. Nat Neurosci 2019. doi:10.1038/s41593-018-0326-7.

64 Levey DF, Stein MB, Wendt FR, Pathak GA, Zhou H, Aslan M et al. Bi-ancestral depression GWAS in the Million Veteran Program and meta-analysis in >1.2 million individuals highlight new therapeutic directions. Nat Neurosci 2021. doi:10.1038/s41593-021-00860-2.

65 Karlsson Linnér R, Biroli P, Kong E, Meddens SFW, Wedow R, Fontana MA et al. Genome-wide association analyses of risk tolerance and risky behaviors in over 1 million individuals identify hundreds of loci and shared genetic influences. Nat Genet 2019. doi:10.1038/s41588-018-0309-3.

66 Grotzinger AD, Rhemtulla M, de Vlaming R, Ritchie SJ, Mallard TT, Hill WD et al. Genomic structural equation modelling provides insights into the multivariate genetic architecture of complex traits. Nat Hum Behav 2019. doi:10.1038/s41562-019-0566-x.

67 O’Connor LJ, Price AL. Distinguishing genetic correlation from causation across 52 diseases and complex traits. Nat Genet 2018. doi:10.1038/s41588-018-0255-0.

68 Jia H, Zack MM, Gottesman II, Thompson WW. Associations of Smoking, Physical Inactivity, Heavy Drinking, and Obesity with Quality-Adjusted Life Expectancy among US Adults with Depression. Value Heal 2018. doi:10.1016/j.jval.2017.08.002.

69 Metse AP, Clinton-Mcharg T, Skinner E, Yogaraj Y, Colyvas K, Bowman J. Associations between suboptimal sleep and smoking, poor nutrition, harmful alcohol consumption and inadequate physical activity (‘snap risks’): A comparison of people with and without a mental health condition in an australian community survey. Int J Environ Res Public Health 2021. doi:10.3390/ijerph18115946.

70 R Core Team (2021). R: A Language and Environment for Statistical Computing. R Found. Stat. Comput. 2021.

71 Baniqued PL, Gallen CL, Voss MW, Burzynska AZ, Wong CN, Cooke GE et al. Brain network modularity predicts exercise-related executive function gains in older adults. Front Aging Neurosci 2018. doi:10.3389/fnagi.2017.00426.

72 Chaddock-Heyman L, Weng TB, Kienzler C, Weisshappel R, Drollette ES, Raine LB et al. Brain Network Modularity Predicts Improvements in Cognitive and Scholastic Performance in Children Involved in a Physical Activity Intervention. Front Hum Neurosci 2020. doi:10.3389/fnhum.2020.00346.

73 Smallwood J, Bernhardt BC, Leech R, Bzdok D, Jefferies E, Margulies DS. The default mode network in cognition: a topographical perspective. Nat. Rev. Neurosci. 2021. doi:10.1038/s41583-021-00474-4.

74 Wang S, Tepfer LJ, Taren AA, Smith D V. Functional parcellation of the default mode network: a large-scale meta-analysis. Sci Rep 2020. doi:10.1038/s41598-020-72317-8.

75 Mirza SS, Ikram MA, Bos D, Mihaescu R, Hofman A, Tiemeier H. Mild cognitive impairment and risk of depression and anxiety: A population-based study. Alzheimer’s Dement 2017. doi:10.1016/j.jalz.2016.06.2361.

76 Perini G, Ramusino MC, Sinforiani E, Bernini S, Petrachi R, Costa A. Cognitive impairment in depression: Recent advances and novel treatments. Neuropsychiatr Dis Treat 2019. doi:10.2147/NDT.S199746.

77 Rock PL, Roiser JP, Riedel WJ, Blackwell AD. Cognitive impairment in depression: A systematic review and meta-analysis. Psychol. Med. 2014. doi:10.1017/S0033291713002535.

78 Delgado MR, Nearing KI, LeDoux JE, Phelps EA. Neural Circuitry Underlying the Regulation of Conditioned Fear and Its Relation to Extinction. Neuron 2008. doi:10.1016/j.neuron.2008.06.029.

79 Morawetz C, Bode S, Baudewig J, Kirilina E, Heekeren HR. Changes in Effective Connectivity Between Dorsal and Ventral Prefrontal Regions Moderate Emotion Regulation. Cereb Cortex 2016. doi:10.1093/cercor/bhv005.

80 Ochsner KN, Gross JJ. The cognitive control of emotion. Trends Cogn. Sci. 2005. doi:10.1016/j.tics.2005.03.010.

81 Koenigs M, Grafman J. The functional neuroanatomy of depression: Distinct roles for ventromedial and dorsolateral prefrontal cortex. Behav. Brain Res. 2009. doi:10.1016/j.bbr.2009.03.004.

82 Nejati V, Majdi R, Salehinejad MA, Nitsche MA. The role of dorsolateral and ventromedial prefrontal cortex in the processing of emotional dimensions. Sci Rep 2021. doi:10.1038/s41598-021-81454-7.

83 Yanagisawa H, Dan I, Tsuzuki D, Kato M, Okamoto M, Kyutoku Y et al. Acute moderate exercise elicits increased dorsolateral prefrontal activation and improves cognitive performance with Stroop test. Neuroimage 2010. doi:10.1016/j.neuroimage.2009.12.023.

84 Li MY, Huang MM, Li SZ, Tao J, Zheng GH, Chen LD. The effects of aerobic exercise on the structure and function of DMN-related brain regions: a systematic review. Int. J. Neurosci. 2017. doi:10.1080/00207454.2016.1212855.

85 Tao J, Liu J, Egorova N, Chen X, Sun S, Xue X et al. Increased hippocampus-medial prefrontal cortex resting-state functional connectivity and memory function after Tai Chi Chuan practice in elder adults. Front Aging Neurosci 2016. doi:10.3389/fnagi.2016.00025.

86 De Hert M, Correll CU, Bobes J, Cetkovich-Bakmas M, Cohen DAN, Asai I et al. Physical illness in patients with severe mental disorders. I. Prevalence, impact of medications and disparities in health care. World Psychiatry. 2011. doi:10.1002/j.2051-5545.2011.tb00014.x.

87 Fang H, Tu S, Sheng J, Shao A. Depression in sleep disturbance: A review on a bidirectional relationship, mechanisms and treatment. J. Cell. Mol. Med. 2019. doi:10.1111/jcmm.14170.

88 Cox RC, Olatunji BO. A systematic review of sleep disturbance in anxiety and related disorders. J. Anxiety Disord. 2016. doi:10.1016/j.janxdis.2015.12.001.

89 Cheng W, Rolls E, Gong W, Du J, Zhang J, Zhang XY et al. Sleep duration, brain structure, and psychiatric and cognitive problems in children. Mol Psychiatry 2021. doi:10.1038/s41380-020-0663-2.

90 Brenes GA, Miller ME, Stanley MA, Williamson JD, Knudson M, McCall WV. Insomnia in older adults with generalized anxiety disorder. Am J Geriatr Psychiatry 2009. doi:10.1097/JGP.0b013e3181987747.

91 Lippman S, Gardener H, Rundek T, Seixas A, Elkind MSV, Sacco RL et al. Short sleep is associated with more depressive symptoms in a multi-ethnic cohort of older adults. Sleep Med 2017. doi:10.1016/j.sleep.2017.09.019.

92 Li Y, Wu Y, Zhai L, Wang T, Sun Y, Zhang D. Longitudinal Association of Sleep Duration with Depressive Symptoms among Middle-aged and Older Chinese. Sci Rep 2017. doi:10.1038/s41598-017-12182-0.

